# TMS–EEG Reveals Distinct Cortical Signatures in Non-Fluent PPA

**DOI:** 10.64898/2025.12.15.25342283

**Authors:** Francesco Lomi, Sofia Santicioli, Silvia Casarotto, Ali Jafarov, Gabriel Hassan, Alessandro Giannotta, Carmelo L. Smeralda, Francesco Neri, Giulia Stramucci, Adriano Scoccia, Lucia Monti, Alberto Benelli, Sonia Padiglioni, Andrea Mignarri, Salvatore Mazzeo, Valentina Moschini, Carmen Morinelli, Pantelis Lioumis, Valentina Bessi, Simone Rossi, the Alzheimer’s Disease Neuroimaging Initiative

## Abstract

Primary Progressive Aphasias (PPA) are a group of neurodegenerative disorders characterized by the gradual decline of language abilities. They are typically divided into three major clinical variants: the non-fluent (nfvPPA), the semantic (svPPA) and the logopenic (lvPPA) variant. Even with an extensive clinical examination, a correct differential diagnosis among variants can be difficult due to the overlapping of dysfunctional language features. In this context, the combination of Transcranial Magnetic Stimulation and Electroencephalography (i.e., TMS–EEG) could extend our understanding of nfvPPA pathophysiology, given the possibility to non-invasively and directly measure cortical reactivity of brain speech networks to external perturbations. Twenty PPA patients (7 nfvPPA, 13 lvPPA) and 8 elderly controls underwent a TMS–EEG session targeting the left dorsal premotor cortex (Brodmann area 6). A subset of 9 patients (8 lvPPA, 1 nfvPPA) were additionally stimulated in the right homologous region. We automatically detected the EEG channel under the stimulator with the highest peak-to-peak amplitude of the early TMS-evoked response and computed the following measures: (i) natural frequency; (ii) normalized evoked spectral power in the alpha, low-beta, high-beta and gamma range. Non-fluent PPA patients showed a slower and simplified TMS-evoked response as compared to healthy elderly subjects, namely a reduction in high-beta power and natural frequency coupled with higher low frequencies (i.e., alpha) intrusion. No significant differences were detected between lvPPA and controls or nfvPPA and lvPPA. The speech rate was positively correlated with TMS–EEG measures (the high-beta power and the natural frequency). Furthermore, compared to the left side, the stimulation of the right hemisphere elicited TMS-evoked responses with higher natural frequency and high-beta power in both lvPPA and nfvPPA patients. This study first shows that TMS–EEG may provide useful neurophysiological biomarkers for characterizing the nfvPPA variant and monitor disease progression across variants. These findings might be employed in the future to stratify patients and eventually inform the application of variant-specific stimulation protocols tailored to individual neurophysiological profiles.

## Introduction

Primary Progressive Aphasias (PPA) represents a cluster of rare neurodegenerative syndromes caused by the deterioration of brain language networks. The main diagnostic criterion for PPA consists of a prominent and isolated language deficit during the initial phase of the disease^1^. Three main PPA variants have been described in literature: non-fluent/agrammatic (nfvPPA), logopenic (lvPPA) and semantic (svPPA). The non-fluent phenotype is characterized by the difficulty in articulating speech (i.e., speech apraxia), which leads to slow speech rate and sound distortions, and/or the deficit in grammatical knowledge (i.e., agrammatism). The cortical brain regions with prominent atrophy in nfvPPA encompass the left inferior frontal, insular, premotor, and supplementary motor regions^2,3^. On the other hand, lvPPA manifests as impaired single-word retrieval and impaired sentence repetition, and early neuropathological changes occur in temporoparietal regions, mainly in the left hemisphere^4^. Finally, svPPA presents instead with the loss of semantic knowledge that manifests with anomia and deficits in single-word comprehension, due to the selective involvement of the left anterior temporal lobe^5^. The PPA subtypes are probabilistically associated with distinct neuropathological substrates: while nfvPPA and svPPA cases are typically associated with frontotemporal lobar degeneration (FTLD), lvPPA is mostly caused by Alzheimer’s disease (AD) pathology^6^.

The overall prevalence of FTLD has recently been estimated, based on a meta-analysis, to be about 10 per 100,000 people^7^. Within FTLD, the prevalence is approximately 3 per 100,000 for nfvPPA and about 1 per 100,000 for svPPA^7^. In a population-based study, the prevalence of lvPPA was 98 per 1,000,000 individuals in the 55–59 age group and 228 per 1,000,000 individuals in the 60–64 age group^8^. Despite being relatively rare conditions in the general population, their typical early onset represents a significant burden on society^9,10^.

Unfortunately, there is no effective pharmacological treatment for PPA at the present stage, and the current approaches available aim to solely counteract disease progression. Speech and Language Therapy (SLT) is commonly used to strengthen the patient’s residual abilities and implement compensatory strategies. Non-invasive Bran Stimulation (NIBS) techniques, as transcranial direct current stimulation (tDCS) or repetitive transcranial magnetic stimulation (rTMS), could provide significant boost of SLT effects, but further studies are needed to confirm these preliminary results^11^. Although there are no resolutive interventions currently available, research is underway to optimize PPA diagnosis and management. Effective management clearly depends on the accurate identification of clinical variants, which can be challenging in PPA due to overlapping clinical manifestations in the language domain^12^. For example, the naming impairment can be found in each variant, although its cause is variant-specific. Due to difficulty in word retrieval, especially in lvPPA, the resulting speech can be slowed in a similar way to what is usually seen in nfvPPA. This heterogeneity of clinical presentations leads to unclassified or mixed PPA cases^13^. Extensive neurological, neuropsychological, and speech-language assessments are typically necessary to correctly diagnose each variant, often leading to fatigue and frustration for the individuals undergoing testing.

Considering its high cost-effectiveness, widespread availability, and low invasiveness, resting-state Electroencephalography (i.e. EEG) has been explored as a potential source of biomarker for the differentiation of PPA phenotypes^14–17^, with less than conclusive results. In this context, the combination between TMS and Electroencephalography (TMS–EEG), that enables a reproducible, non-invasive, spatially and temporally precise investigation of cortical reactivity, could offer new insights into the neuropathology of PPA. Unlike traditional TMS, which is limited to the motor cortex via Motor Evoked Potentials (MEPs) readout, TMS–EEG allows the assessment of local reactivity and connectivity even across non-motor networks^18^. Compared to EEG alone, it provides higher spatial resolution and the possibility to infer causal brain–behavior relationships.

In this regard, the dorsal premotor cortex (PMd) in the left hemisphere represents a plausible candidate for TMS–EEG targeting in PPA. Above all, premotor regions play a crucial role in speech perception and production. Together with inferior frontal and inferior parietal regions they form the “dorsal stream” of the language network responsible for the mapping from phonological to articulatory representations^19^. According to the highly influential DIVA (*Directions Into Velocities of Articulators*) model^20^, premotor regions underpin the speech sound representations that trigger the feedforward system to activate motor programs and produce speech sounds^21^. In nfvPPA, atrophy mainly spreads from posterior inferior frontal to anatomically and functionally connected regions, like PMd and SMA via frontal aslant tract^3^. The functional significance of these regions is also confirmed by correlational evidence between the degree of atrophy and articulation rate in nfvPPA^22^.

Due to its medial location within the borders of the superior frontal gyrus, the PMd is less prone to TMS-induced muscle artifacts, which more commonly affect lateral frontal regions such as the dorsolateral prefrontal cortex (dlPFC) or the inferior frontal gyrus (IFG)^23^. As a result, TMS–EEG recordings obtained from this site typically exhibit cleaner signals that require only minimal preprocessing.

So far, no prior study has applied TMS–EEG in the attempt to characterize PPA phenotypes. Here, we hypothesized that:

a. the stimulation of the left PMd would reveal abnormal TMS–EEG patterns in nfvPPA compared to controls, given the specific involvement of this region and BA6 in general in this variant; and
b. the stimulation of PMd would elicit different TMS-evoked responses when targeting the left versus the right hemisphere in PPA patients, reflecting their asymmetric pathological involvement.

## Materials and methods

### Patients and clinical evaluation

We longitudinally included patients referred to the center for Cognitive Disturbance of the Neurology Unit of Careggi at the University Hospital of Florence and to the Neurology Unit at University of Siena that fulfilled the current consensus criteria for PPA defined by Gorno-Tempini and colleagues (2011)^1^. Twentyfour patients clinically diagnosed with PPA (14 lvPPA, 10 nfvPPA) participated in the study. All patients or their caregivers provided written informed consent before study inclusion. Eight age-matched healthy controls (HC) were also recruited after informed consent and underwent the same TMS–EEG investigation of PPA patients. They were clinically evaluated to confirm preserved cognitive functions within a normal range. The study was conducted according to the guidelines of the Declaration of Helsinki and was approved by the local ethical committee (protocol code: Brainsight 21/24).

Patients were classified into PPA variants by three neurologists (VB and SM) and three neuropsychologists (SP, VM, CM) with expertise in cognitive disorders, according to the current diagnostic criteria of PPA^1^. Prior established exclusion criteria were: i) non-native Italian speakers; ii) patients with a history of head injury, current neurological and/or systemic disease, or substance use disorder; iii) right predominant PPA; and iv) patients with severe language impairment that did not allow neuropsychological evaluation. All participants underwent family and clinical history collection, neurological examination, extensive neuropsychological investigation, including speech and language testing.

### Neuropsychological assessment

All PPA patients were evaluated by an extensive neuropsychological battery consisting of global measurements (MMSE) and specific tasks exploring each cognitive function. Details about the tests employed have been previously described elsewhere^24^.

Speech fluency was assessed using a standardized picture description task. Participants were presented with the validated “Cookie Theft Picture” from the Boston Diagnostic Aphasia Examination^25^ and were instructed to describe the scene in their own words, as if the examiner could not see the picture. Speech samples were audio-recorded and subsequently transcribed by a trained neuropsychologist with experience in linguistic data collection (FL).

Fluency (words/sec) was quantified by calculating the ratio between the total number of words produced and the total duration of the participant’s speech. Examiner prompts and meta-linguistic comments (e.g., remarks about the task itself or about one’s own performance) were excluded from speech total duration. In the word count, false starts (i.e., initial attempts at word production that were not completed) were not included.

### TMS–EEG procedure

In each patient and control subject, structural MRI (3D T1-weighted image) was collected to allow for neuronavigation with TMS. In addition, T2-weighted images were acquired in patients and controls to exclude ischemic or degenerative lesions. Three healthy controls were navigated on the Colin 27 template because of missing MRI data^26^. The TMS target was individually selected over the left posterior portion of the superior frontal gyrus (i.e., left dorsal premotor cortex) and the Brodmann brain atlas embedded in the neuronavigation system (BrainNET, EBneuro Ltd, Florence, Italy) was used to confirm that this target corresponded to the Brodmann Area 6 (i.e., BA6).

During the experiment, participants sat comfortably on a reclining chair in a quiet room. Biphasic TMS pulses were delivered with a figure-of eight coil placed tangentially to the head and connected to an STM9000 stimulator (Ates-EBNeuro). The neuronavigation system was used to monitor coil position in real time. EEG was recorded with a TMS-compatible amplifier (g.HIAMP, g.tec Medical Engineering GmbH, Austria) equipped with a 64-channel cap using a standard montage and passive C-ring-shaped electrodes. Impedance at all electrodes was kept below 5kΩ throughout the recording session. Reference and ground electrodes were located on the forehead above the frontal sinuses. EEG data were acquired in MATLAB (v2020, MathWorks, Natick, MA) via Simulink interface at a sampling rate of 4800 Hz.

The occurrence of evoked auditory artifacts was minimized using a TMS-specific noise masking track built within a dedicated toolbox (TAAC)^27^ and played with noise-cancelation in-ear headphones (Shure SE215). The quality of the TMS-evoked EEG response was ensured by employing an online, real-time, trial-to-trial graphical interface displaying the signal in average reference^28^. First, the selected target was stimulated orthogonally to the midline in light of evidence showing that evoked response is maximized with this orientation^29^. Then, EEG responses to 20 single TMS pulses were averaged and visually inspected. The individual optimal stimulation parameters were then determined in real time based on the observed response. Specifically, we sought for a peak-to-peak amplitude of ∼8 μV in the 20-60 ms time window on the channel nearest to the coil (e.g., FC1), while minimizing decay or muscle-related artifacts. Stimulus intensity and/or coil orientation were slightly adjusted if needed. Otherwise, if no clear response was detected, also the TMS target was slightly adjusted. The real-time inspection was then repeated until the above criterion was reached. Each recorded TMS–EEG session consisted of 210 single pulses with a jittered interstimulus interval (3-5 sec with 100 ms step). Three nfvPPA patients were excluded from final sample because of small-to-absent local TMS-evoked responses due to severe cortical atrophy (see Supplementary Figure 2), whereas one lvPPA was excluded because of insufficient level of vigilance during the experiment.

In a subset of patients (*n* = 9, 1 nfvPPA and 8 lvPPA), the contralateral PMd in the right hemisphere was also stimulated during the same session. Starting from the exact corresponding location in the right hemisphere, mirroring the target identified in the left hemisphere, the coil position was adjusted slightly until a peak-to-peak amplitude of approximately 8 μV was obtained in the 20–100 ms time window on the electrode closest to the coil (e.g., FC2). The stimulation intensity was kept identical to that used for the left hemisphere, and the neuronavigation system ensured consistent coil orientation and angle across hemispheres.

### TMS–EEG data preprocessing and analysis

Data pre-processing was performed using customized algorithms based on the EEGLAB toolbox running on MATLAB (v2023)^30^. EEG data was first segmented into trials of 1600 ms around the TMS pulse. To remove the pulse artifact, we replaced an 8-ms long time-window around the pulse with an equivalent amount of baseline. After high-pass (1 Hz, Butterworth, 3rd order) and notch (49-51 Hz, 3^rd^ order) filtering applied to continuous data, bad channels and epochs were selected; then, baseline correction (−800.0 −0.2 ms) and re-reference to the average were applied. ICA (EEGLAB runica function) was subsequently applied to remove residual artifacts of ocular or muscular origin. The dataset was then low-pass filtered (45 Hz, Butterworth, 3rd order), downsampled to 1/5 of the original frequency (960 Hz) and re-segmented to the window of ±600 ms surrounding the TMS pulse. Lastly, signals from bad channels were interpolated using spherical splines.

Then, we automatically identified the EEG channel under the stimulator as the one with the highest peak-to-peak amplitude of the early TMS-evoked response (< 80 ms). We applied time-frequency analysis on this channel by applying the Stockwell transform (ST), i.e., a method that combines Short Time Fourier Transform with signal filtering by frequency-dependent Gaussian scaling windows^31^. The width of the Gaussian window was set at 0.8 for a better temporal resolution. Time-frequency power spectra were baseline-corrected by subtracting the average pre-stimulus power (from −500 to −100 ms) from each frequency bin. Then, normalized evoked power was computed by cumulating within a 20-200 ms post-TMS window in the alpha (8-13 Hz), low-beta (13-20 Hz), high-beta (20-30 Hz) and gamma (30-45 Hz) ranges. The natural frequency was defined as the frequency *f* at which the evoked power reached its maximum value (peak frequency)^32^. The same measures were computed also by considering the same EEG channel (i.e., FC1) for all subjects (see Supplementary Results). To verify that the applied stimulation intensity was sufficient to evoke a reliable response in patients and healthy controls, we computed the induced E-field intensity over the TMS hotspot with SimNIBS software (version 4.1)^33^. It has been indeed demonstrated that an induced field between 90 and 130 V/m provides TMS–EEG responses with good signal-to-noise ratio^32^. Simulations in SimNIBS were optimized implementing brain surfaces generated on FreeSurfer (v8.0.0)^34^.

### VBM analysis

Magnetic Resonance Images (MRIs) were utilized to conduct voxel-based morphometry (VBM) analysis to identify regions of atrophy (i.e., morphometric differences) across all PPA variants in comparison to HC^35^. For this analysis, we employed the Computational Anatomy Toolbox (CAT12), an extension of the Statistical Parametric Mapping (SPM, Wellcome Department of Cognitive Neurology, v25) software and operated within MATLAB (v2023)^36,37^.

Due to the suboptimal quality of the MRI scans in the HC group, additional data used in the preparation of this article were obtained from the Alzheimer’s Disease Neuroimaging Initiative (ADNI) database (adni.loni.usc.edu). The ADNI was launched in 2003 as a public-private partnership, led by Principal Investigator Michael W. Weiner, MD. The primary goal of ADNI has been to test whether serial magnetic resonance imaging (MRI), positron emission tomography (PET), other biological markers, and clinical and neuropsychological assessment can be combined to measure the progression of mild cognitive impairment (MCI) and early Alzheimer’s disease (AD).

Three-dimensional T1-weighted Magnetization Prepared Rapid Gradient Echo (MPRAGE) DICOM images of 67 cognitively normal (CN) controls (36 females, 31 males) were downloaded from the ADNI1: Screening project. Selected data were acquired using 1.5 Tesla MRI scanners from Siemens Systems (Siemens Healthineers, Germany) and filtered by age, encompassing individuals aged 60 to 80 years (mean of 73.96 yo). PPA patients (2 nfvPPA, 3 lvPPA) with anisotropic MRI scans were also excluded from VBM analysis, as their images did not meet the quality criteria for statistical analysis.

We preprocessed 3D T1-weighted MRI scans using the CAT12 manual, which includes the default segmentation pipeline, quality control measures and spatial smoothing procedure^26^. We computed the total intracranial volume (TIV) to normalize the modulated gray matter (GM) tissue images for variability in brain size and volume, employing an analysis of covariance (ANCOVA) approach. We ensured the homogeneity of samples by reviewing all images. To enhance the statistical robustness of our group comparisons, we applied an 8×8×8 mm full width at half maximum (FWHM) Gaussian smoothing kernel to the data. The volumetric statistical results for each PPA group were saved as thresholded brain masks and visualized by overlaying these masks on a reference brain template using BrainNet Viewer^38^.

## Statistical Analysis

Between-subjects analyses were performed to compare demographic, clinical, and TMS parameters, as well as neurophysiological measures obtained from TMS–EEG (including natural frequency and spectral power in different frequency bands) across the three groups: nfvPPA, lvPPA and HC. One-way analyses of variance (ANOVA) were performed for continuous variables when assumptions of homoscedasticity and normality of residuals were met; otherwise, the non-parametric Kruskal-Wallis test was applied. Post-hoc comparisons were adjusted to control for inflated type I error. Categorical variables (e.g., sex) were analyzed using the Chi-squared test.

Correlations between variables were examined using Pearson’s correlation coefficient. Normality of each variable was assessed with the Shapiro–Wilk test to ensure the appropriateness of parametric correlation analysis.

Within-subjects analyses were conducted to compare responses between the left and right hemispheres. Spectral power across frequency bands (alpha, beta1, beta2, gamma) was analyzed using a linear mixed-effects model including Hemisphere, Band, and their interaction as fixed factors, with Subject modeled as a random intercept to account for repeated measures. A singular fit warning indicated that the variance of the random intercept was estimated close to zero, likely due to the small sample size. Post-hoc pairwise contrasts between hemispheres within each band were computed using estimated marginal means. Natural frequency, E-field, TMS coordinates and peak-to-peak differences between hemispheres were assessed using paired-sample t-tests after confirming normality of within-subject differences, otherwise Wilcoxon test was used.

For VBM analysis, a two-sample t-test model was used to determine the significant difference between the means of the PPA variants and the HC. The final models were calculated and compared using an uncorrected threshold of *p* < 0.001 with an empirically determined extended threshold, and false discovery rate (FDR) correction with a significance level (*p* < 0.01).

## Data availability

The data that support the findings of this study and all custom-written analysis codes are available from the corresponding author upon reasonable request.

## Results

### Demographic and clinical characteristics

Demographic and clinical characteristics of final sample (13 lvPPA, 7 nfvPPA, 8 HC) are reported in Table 1. No differences in sex, age, or education years were found. Moreover, the PPA groups did not differ significantly in terms of disease duration. There was instead a significant difference in MMSE score, and a significantly lower MMSE for lvPPA than healthy controls at post-hoc test. As expected, the speech rate differed between PPA groups (*F*(1,18) = 24.41, *P* < 0.001): nfvPPA was associated with a lower speech rate as compared with logopenic patients (*P* < 0.001).

**Table 1.** Mean demographics and clinical characteristics of included sample nfvPPA = non-fluent variant PPA, lvPPA = logopenic variant PPA, HC = healthy controls, MMSE = Mini Mental State Examination, SD = standard deviation.

|  | **nfvPPA (*n* = 7)** | **lvPPA (*n* = 13)** | **HC (*n* = 8)** |  |
| --- | --- | --- | --- | --- |
| Age, years, mean (SD) | 74.6 (3.55) | 69.31 (7.36) | 66 (9.56) | *P* = 0.177 |
| Sex, females, *n* (%) | 1 (14%) | 8 (62%) | 3 (60%) | *P* = 0.118 |
| Education, years, mean (SD) | 14.9 (4.71) | 13.2 (3.48) | 14.1 (7.06) | *P* = 0.648 |
| Disease duration, mean (SD) | 5.00 (1.73) | 3.85 (2.12) | - | *P* = 0.233 |
| MMSE, corrected score, mean (SD) | 27.1 (5.93) | 23.4 (3.70) | 29.3 (1.41) | *P* < 0.01 |
| Speech rate, words/sec, mean (SD) | 0.58 (0.24) | 1.22 (0.29) | - | *P* < 0.001 |

### TMS–EEG parameters

TMS–EEG parameters of the study population are reported in Supplementary Table 1. On average, all groups were stimulated with very similar intensities, both in terms of stimulator output percentage and maximum induced E-field. The maximal E-field strength was in the range considered as optimal in literature to evoke genuine TMS responses (see Methods). Importantly, the MNI coordinates for the TMS hotspot did not significantly differ across groups. There were also no significant differences across groups in terms of the electrode of interest automatically extracted and used for the analysis and peak-to-peak amplitude over that channel.

In the comparison between left and right hemispheres (Supplementary Table 2), no significant differences emerged for target coordinates, induced E-field intensity, or peak-to-peak amplitudes.

### Grey matter atrophy in PPA variants

Figure 1 shows the results of VBM analysis with two different statistical thresholds (uncorrected *P* < 0.001 and FDR < 0.01). Patients with nfvPPA were associated with left posterior-inferior frontal and middle frontal/premotor atrophy, while lvPPA patients showed a more widespread and less asymmetric pattern of atrophy involving temporal, parietal and frontal cortices.

**Figure 1.**
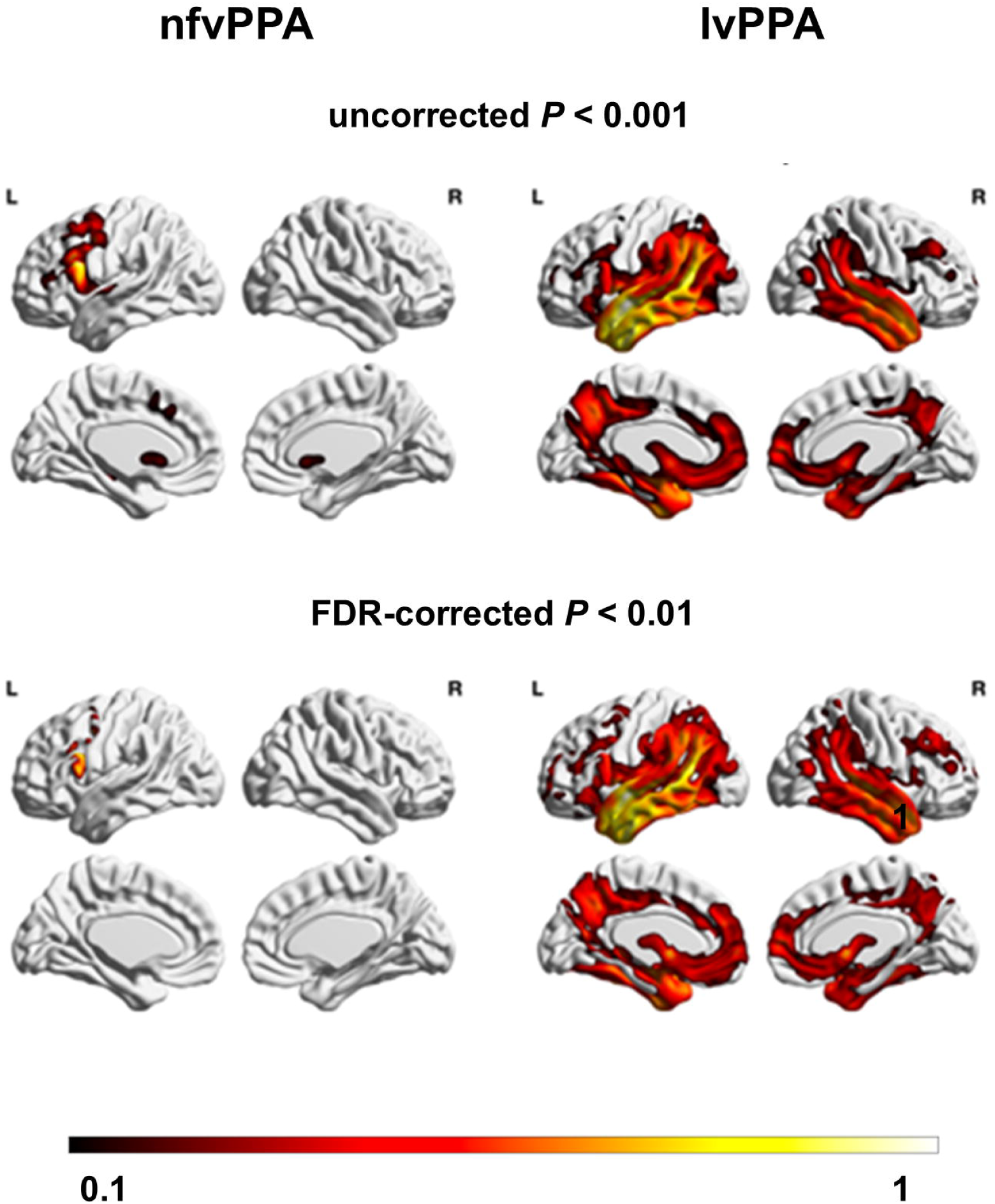
Atrophy patterns in nfvPPA and lvPPA. Regions of atrophy in nfvPPA and lvPPA patients compared to controls, either uncorrected for multiple comparisons at *P* < 0.001 (above) or after correction for multiple comparisons using false discovery rate (FDR) correction at *P* < 0.01 (below). The colour scale refers to the magnitude of the two-sample t-statistic at each voxel.

### Characterization of TMS-evoked cortical activity

#### Non-fluent PPA is associated with a reduction in premotor high-beta power and natural frequency in the left hemisphere

Figure 2 shows the TMS hotspot coordinates in MNI space for all subjects (Panel A) and the grand average TEP for each group with the respective topographical plots at relevant timepoints (Panel B-D). Both in PPA patients and in HC, single-pulse TMS evoked a clear positive peak in the first 30 ms in the area under the coil. A second peak of negative polarity occurred on average between 40 and 70 ms over the same regions, likely reflecting hyperpolarization of the same neuronal circuits. In the nfvPPA group, the first negative peak was significantly delayed compared to controls (X^2^(2) = 7.53, *P* = 0.023; *P* = 0.013). Non-fluent PPA patients also showed an increased latency of the second positive peak compared to HC (*F*(2,25) = 5.28, *P* = 0.012; *P* = 0.009). No post-hoc differences were detected between lvPPA and HC or between lvPPA and nfvPPA.

**Figure 2.**
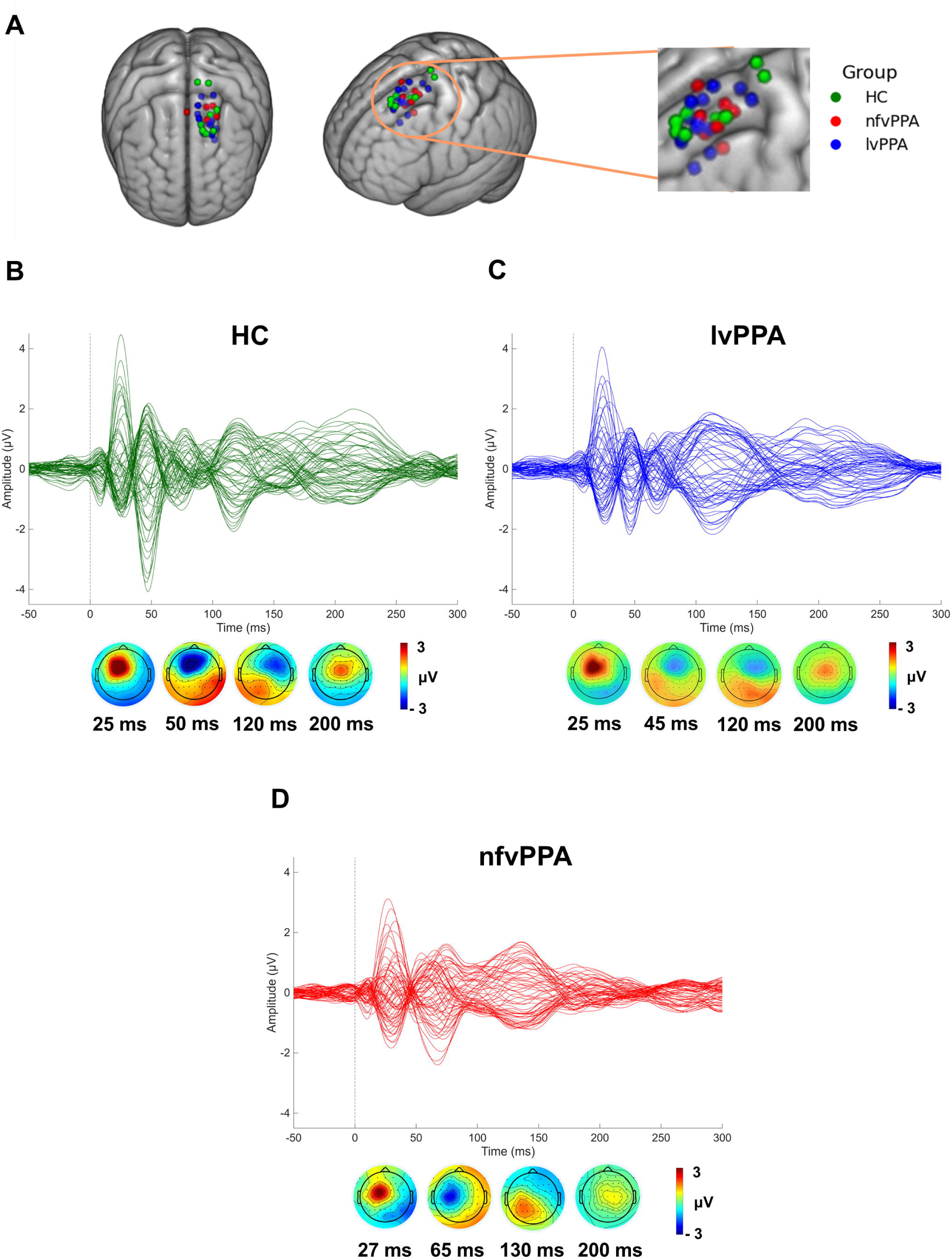
TMS hotspots and evoked potentials. (**A**) TMS hotspot coordinates in MNI space for all subjects. (**B-D**) Grand-average butterfly plots of TMS-evoked potentials (TEPs) for each group with the respective topographical plots at relevant timepoints.

Regarding TMS-evoked relative power, in the high-beta range (21-30 Hz) there was a statically significant difference between groups (*F*(2,25) = 5.85, *P* = 0.008), with nfvPPA showing a significantly lower power spectral density than controls (*P* = 0.006) (Figure 3A). A significant effect was also evident in the alpha band (X^2^(2) = 6.77, *P* = 0.034), where nfvPPA showed higher alpha power than HC (*P* = 0.042).

**Figure 3.**
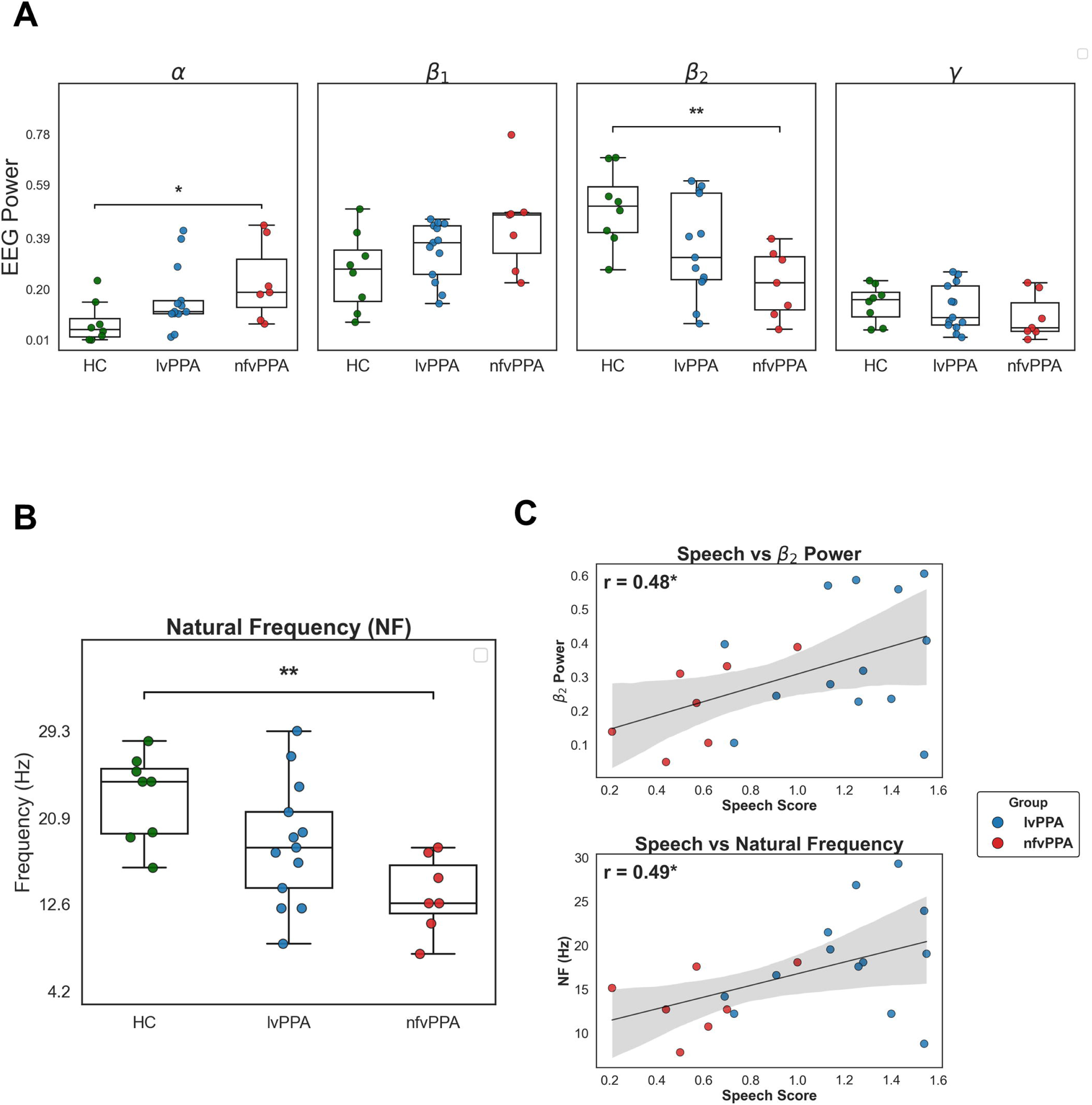
TMS–EEG spectral measures and clinical correlations. (**A**) Box-plots of evoked spectral power in each frequency band for each group. (**B**) Box-plots of natural frequency values for each group. (**C**) Linear relations between the speech rate at clinical evaluation and each TMS–EEG outcome: evoked high-beta power (above) and natural frequency (below). Asterisks indicate significant group differences or correlations (*P* < 0.05; *P* < 0.01; *P* < 0.001).

The shift in power spectral density toward lower frequencies in the nfvPPA group was also evident when analyzing natural frequencies (*F*(2,25) = 6.58, *P* = 0.005), where nfvPPA patients showed a significantly lower natural frequency than healthy subjects (*P* = 0.004) (Figure 3B; see also Figure 4 for representative subjects). No significant differences emerged in terms of natural frequency between nfvPPA and lvPPA (*P* = 0.113), and lvPPA and HCs (*P* = 0.133) at post-hoc tests. No significant group differences were detected in the low-beta (*F*(2,25) = 3.01, *P* = 0.068) and gamma (X^2^(2) = 1.99, *P* = 0.370) power.

**Figure 4.**
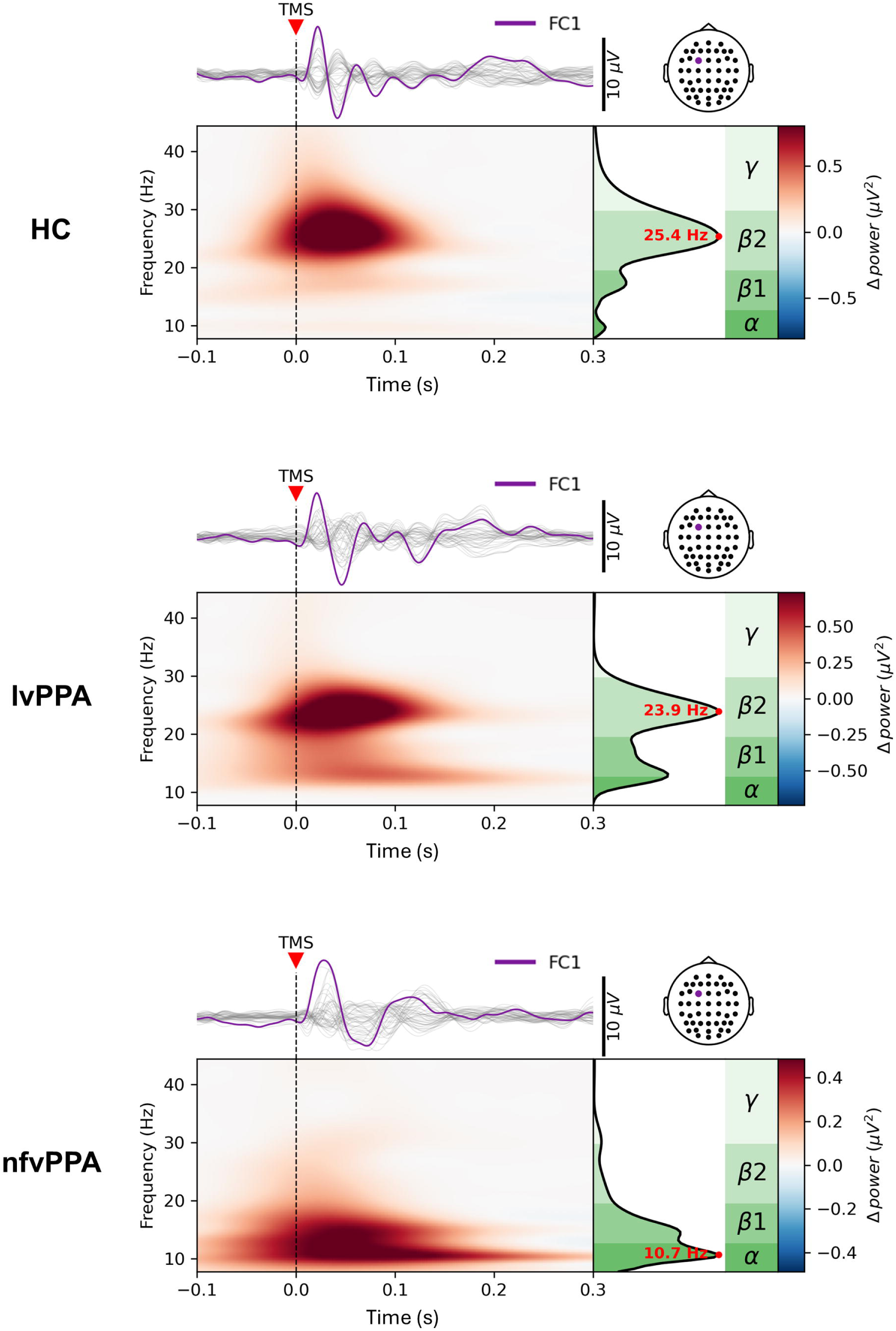
Representative TMS–EEG responses. TMS–EEG responses of three representative subjects, one per group. Each panel shows the butterfly plot, the time-frequency maps of the evoked spectral power and the respective natural frequency over the electrode of interest.

#### TMS–EEG outcomes correlate with speech rate across variants

We then performed correlation analyses between TMS–EEG outcomes and speech rate (words/sec) in the PPA cohort (Figure 3C). We obtained a significant result correlating speech rate with the power in the high-beta range (*r* = 0.48, *P* = 0.031), but not in the other frequency bands. Also the natural frequency correlated significantly with speech rate (*r* = 0.49, *P* = 0.030).

#### Interhemispheric differences in high-beta power and natural frequency in PPA

We examined the effects of Hemisphere and Frequency Band on TMS-evoked power using a linear mixed-effects model with Subject as a random intercept. Type III ANOVA revealed a significant main effect of Band (*F*(3,64) = 16.64, *P* < 0.0001), no significant main effect of Hemisphere (*F*(1,64) < 0.01, *P* = 0.999), and a significant Hemisphere × Band interaction (*F*(3,64) = 3.68, *P* = 0.016) (Figure 5; see also Figure 6 for representative subjects).

**Figure 5.**
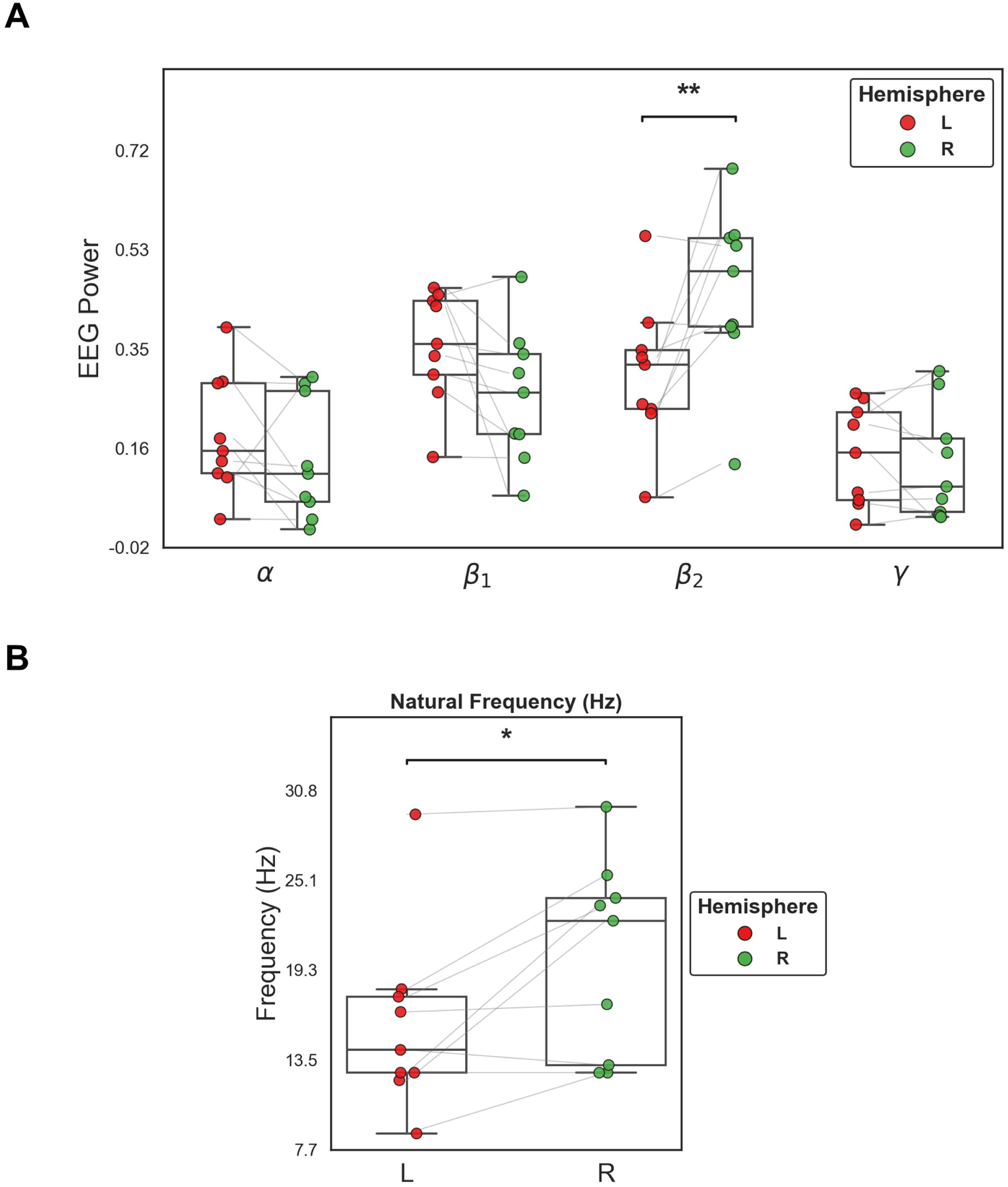
Inter-hemispheric spectral measures. (**A**) Box-plots of evoked spectral power for each frequency band, shown separately for the left and right hemispheres. Individual observations are displayed as coloured points, with colours corresponding to the hemispheric identity as indicated in the legend. (**B**) Box-plots of natural frequency values for the left and right hemispheres, with single-subject data points plotted using the same hemisphere-specific colour coding. Asterisks indicate significant differences between hemispheres (*P* < 0.05; *P* < 0.01; *P* < 0.001).

**Figure 6.**
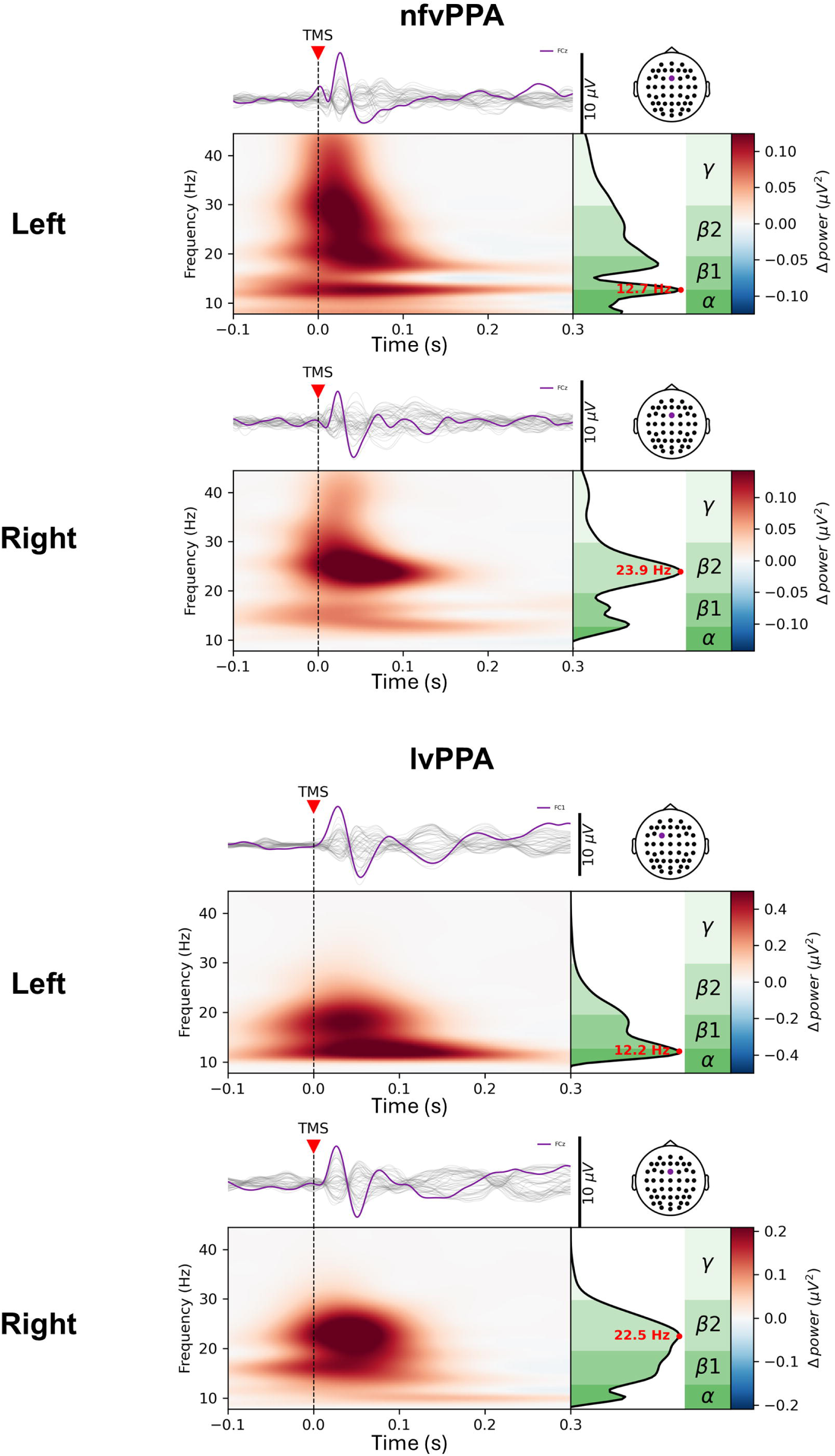
Bilateral representative TMS–EEG responses. TMS–EEG responses from two representative patients (top: nfvPPA; bottom: lvPPA), shown for both the left and right hemispheres. Each panel includes the butterfly plot of TMS-evoked potentials, the time–frequency maps of evoked spectral power, and the corresponding natural frequency measured over the electrode of interest.

Post-hoc pairwise comparisons using estimated marginal means indicated a significant hemispheric difference only in the high-beta band, with lower power in the left hemisphere compared to the right one (estimate = –0.155, *t*(56) = –2.75, *P* = 0.008). No significant hemispheric differences were observed in the alpha (*P* = 0.409), low-beta (*P* = 0.105), or gamma bands (*P* = 0.786).

Natural frequency was significantly lower in the left hemisphere than in the right (*t*(8) = – 2.78, *P* = 0.024) (Fig. 6). The mean hemispheric difference was –3.76 Hz (95% CI: –7.09 to –0.43).

## Discussion

This is the first study employing TMS–EEG with the aim to characterize reactivity of language areas of the frontal cortex in PPA patients. While traditional single-pulse or paired-pulse TMS methods are limited to the study of the primary motor cortex (M1) via Motor Evoked Potentials readout, TMS–EEG provides the unique possibility to non-invasively study functional brain changes in diseases, like PPA, which mostly involve networks outside M1, thus considerably expanding our knowledge on the functional consequences of PPA neuropathology^13^.

In this study we demonstrated that nfvPPA patients show remarkable alterations in the local TMS–EEG response as compared to age-matched HC. These results were observed when stimulating the PMd, located in the left superior frontal gyrus, and were found both on the EEG channel with the maximum peak-to-peak amplitude and on the same channel for all the subjects (i.e., FC1), indicating that the TMS-evoked response was topographically consistent across individuals. Importantly, TMS-evoked EEG measures—specifically high-beta power and natural frequency—were significantly correlated with speech rate, suggesting that these metrics may serve as reliable indicators of the integrity of the speech production network, complementing clinical evaluation. Finally, in a subsample of PPA patients, we observed significant interhemispheric differences in TMS-evoked responses, in line with the asymmetrical progression of neurodegeneration between the two hemispheres (i.e., left dominant more than right, non-dominant).

### Alterations of TMS-evoked EEG responses

The brain response to TMS is the result of polysynaptic activation of corticocortical and corticothalamic networks. While early components of the TEP appear to result from the local activation of the stimulated area, later peaks are thought to be associated with long-range connectivity through axonal signal propagation^39,40^. Common peaks have been identified in sensorimotor regions (e.g., P30, N45, P60, N100)^41^, with high test-retest reproducibility^42^; pharmacological studies have suggested that these might be linked to different neurotransmitter systems (e.g., N45 with GABA_A_ and N100 with GABA_B_)^43^. Furthermore, TMS is thought to synchronize the ongoing pre-existing neural oscillations in a given brain area, allowing to measure the dominant frequency of that specific cortical module^44^. TMS-evoked natural frequencies are region-specific, with a gradient from rostral (with faster oscillations) to occipital, with slower natural frequencies^44^. For premotor regions, the main induced EEG rhythm is in the high-beta/low-gamma range^44,45^.

In the present study, nfvPPA showed globally slower responses than HC when stimulated in the left PMd. In the latter group, TMS-induced responses featured an oscillatory natural frequency in the high-beta range (22.95 ± 4.22 Hz), consistently with the one reported in the literature. By contrast, natural frequency in nfvPPA was shifted toward the low-beta/alpha range (13.53 ± 3.68 Hz). Importantly, in lvPPA the TMS-induced responses were predominantly in the beta band (18.44 ± 5.94 Hz), but substantial variability did not allow to detect significant differences with nfvPPA. This natural frequency reduction in nfvPPA was driven by a breakdown of high-beta power spectral density coupled with augmented alpha power in these patients (see Figures 3A-B and 4).

In this context, structural damage within left BA6 regions in nfvPPA may contribute to the EEG abnormalities we observed. In stroke, increased slow-wave activity—particularly “sleep-like” delta oscillations—in perilesional cortex is well documented^46,47^, with consistent evidence from animal models as well^48^. Altered spectral patterns and impaired oscillatory coherence have been identified as indicators of network dysfunctions underlying specific cognitive deficits across neurological and psychiatric conditions^49,50^. Considering its high cost-effectiveness, widespread availability, and low invasiveness, resting-state EEG has been explored as a potential source of biomarkers for the differentiation of PPA phenotypes. Previous resting-state EEG and Magnetoencephalographic (MEG) studies reported changes in power spectral density in PPA, primarily reflecting an overall slowing of neural oscillations, characterized by reduced relative power in the higher frequency bands, alpha and beta, and increased relative power in the lower frequency bands, delta and theta, compared with controls^14,16,51^. This pattern was observed especially in lvPPA compared with nfvPPA and svPPA^10,11,12^. The different pattern of results reported here has to be attributed to the different kinds of techniques adopted. Indeed, TMS–EEG allows to probe the activity of specific brain regions and network of interest, allowing for causal inference, whereas resting-state EEG and MEG are mainly correlational techniques.

Ranasinghe and colleagues^51^ reported reduced functional connectivity in the beta band in nfvPPA at the source level within the left inferior frontal cortex, which also constitutes the area of peak atrophy in our study. This is consistent with our finding of diminished TMS-evoked high-beta activity in a more dorsal region that nevertheless maintains structural connectivity with inferior frontal areas^52^. Notably, they also described enhanced connectivity in lower-frequencies within the dorsomedial frontal cortex, roughly aligning to our TMS stimulation site, which is also congruent with the relative shift of the TMS-evoked power toward lower frequencies in this study (see Figure 3A). Importantly, we targeted a cortical site that was not significantly affected in our nfvPPA patients (see Figure 1). We excluded three nfvPPA patients where TMS-evoked responses were small-to-absent because of significant global atrophy^53^ (see Supplementary Figure 2).

Therefore, our results are unlikely to reflect a simple epiphenomenon of local atrophy and instead suggest perturbation of a dysfunctional network, potentially driven by structural disconnection among key nodes. In this regard, a crucial role of the thalamus has been postulated in the generation of fast neural oscillations^54^, a region which is involved in FTD phenotypes like nfvPPA^55^. Of note, deficits in GABAergic and glutamatergic neurotransmission have been associated with FTLD pathogenesis and symptomatology^56,57^.

Interestingly, lvPPA patients showed a TMS–EEG response pattern that lies between that of healthy controls and nfvPPA patients and characterized by greater inter-subject variability. This increased heterogeneity may reflect the fact that, despite a similar disease duration, lvPPA tends to progress more rapidly than the other variants^58^. Indeed, MMSE was lower in lvPPA compared to HC. This is supported by our MRI findings, which indicate a more widespread and diffuse pattern of atrophy in lvPPA compared to nfvPPA, as reported also elsewhere^59^. Moreover, we have to consider the inherent heterogeneity of the AD-related PPA spectrum, which includes mixed nfv/lvPPA cases that present with both agrammatism (consistent with nfvPPA) as well as impaired naming and sentences repetition (considered as core criteria for lvPPA)^60^.

Preferential neurodegeneration of the left hemisphere language regions is a common denominator in PPA, even with disease progression^61^. In line with this evidence, we also found that the TMS-evoked responses were higher in high-beta spectral power and natural frequency when stimulating the right compared to the left hemisphere, akin to those found in HC (see Figure 5 and 6). Notably, in a TMS–EEG study, D’Ambrosio and colleagues targeted premotor areas in healthy participants and found no significant hemispheric differences in TMS-evoked spectral features^62^, suggesting that the asymmetry observed in our PPA cohort may reflect disease-related reorganization rather than a non-specific hemispheric imbalance.

### Functional significance of TMS–EEG alterations

As previously mentioned, premotor regions play a crucial role in speech perception and production, representing a key node of the “dorsal stream” of the language network responsible for the mapping from phonological to articulatory representations^19^.

Our results suggest that higher speech rate is associated with stronger and faster intrinsic neural oscillations (see Figure 3C), specifically in the high-beta range, which aligns with the known role of beta sensorimotor rhythms in motor control and timing, including speech production^63^. As observed for externally cued movements, EEG beta oscillations typically desynchronize (i.e., decrease in power) prior to speech onset, probably reflecting feedforward mechanisms that provide the motor plan and the internal representation of intended speech output in corresponding articulatory and sensory cortical regions^64^. The neural sources of this activity are represented by the primary motor and somatosensory cortices (M1/S1), supplementary motor area (SMA), and parietal regions^65^. This is also confirmed by the TMS–EEG literature showing predominant beta rhythm for the sensorimotor system, with lower beta frequencies for parietal regions and higher beta oscillations for premotor regions^44^. Alterations in both the low and high-beta band (i.e., stronger desynchronization) have been reported prior to articulation movements in patients with Parkinson disease^66^ and adults with stuttering^67^ in the EEG literature. Furthermore, weaker pre-movement beta desynchronization correlated with faster speech and hand motor responses in healthy subjects^66^. A recent paper on post-stroke aphasia patients demonstrated that the strength of connectivity in the whole beta band (15-30 Hz) within the affected hemisphere positively correlated with speech fluency^68^.

### Clinical and therapeutic implications

Navigated TMS–EEG has been recently used to characterize and identify adjacent brain circuitries, such as those within BA6^69^. This demonstrates the potential of this method as a non-invasive brain mapping technique capable of delineating the functional profiles of specific brain areas and their associated symptoms^70^.

Non-fluent PPA is characterized by both quantitative (i.e., apraxia of speech) and qualitative (i.e., agrammatism) reductions in speech, making fluency a pivotal diagnostic feature for distinguishing this variant from the others^71^. Given their strong association with speech rate, the alterations we observed in high-beta power and natural frequency over premotor regions may represent a novel objective index of motor speech dysfunction. This neurophysiological signature could aid in refining the characterization of nfvPPA and, more broadly, may hold translational value across multiple speech disorders.

The present findings suggest that TMS–EEG may represent a valuable tool to monitor disease progression in PPA and to provide objective neurophysiological indicators for staging, an area that remains an unmet clinical need^72^. In this context, the interhemispheric differences in high-beta power and natural frequency observed in our cohort could serve as sensitive markers of functional integrity and compensatory mechanisms, potentially aiding in the longitudinal tracking of disease evolution and in the stratification of patients according to disease stage.

### Limitations and future studies

One limitation of this study is the relatively small sample size, which reflects the rarity of these conditions in the general population. Nevertheless, the cognitive profiles and atrophy patterns observed in our PPA cohort are consistent with previously published findings. It is also worth noting that the initial sample size was comparable to that of other neurophysiological studies in PPA^51^, but a minority of patients had to be excluded because their recordings did not meet the minimum criteria for reliable analysis.

As the first study of its kind, these results provide a foundation for future investigations involving larger patient cohorts. An interesting future direction could be to map multiple targets in the language network to neurophysiologically stratify each variant. This multi-focal approach might also inform on the most spared language nodes in each PPA variant, in terms of reactivity and/or connectivity, that might be therefore more prone to the neuromodulation treatment.

## Conclusions

This study provides the first evidence that TMS–EEG can non-invasively characterize functional alterations in the dominant frontal cortex of patients with nfvPPA. By directly probing the left dorsal premotor cortex, we demonstrated that nfvPPA patients exhibit markedly reduced high-beta oscillatory activity and lower natural frequency. These findings align with structural MRI evidence of atrophy in premotor and inferior frontal regions, reinforcing the link between local cortical integrity and electrophysiological reactivity. The observed correlations between TMS–EEG measures and speech rate highlight the clinical relevance of these indices as potential biomarkers of motor speech dysfunction. Importantly, the interhemispheric differences observed both in lvPPA and nfvPPA, showing relatively preserved right-hemisphere reactivity compared to the left, further support the translational potential of TMS–EEG in staging and monitoring PPA.

Despite the discussed limitations, our results demonstrate the feasibility and sensitivity of TMS–EEG for assessing disease-specific neurophysiological signatures beyond the motor cortex. Future studies with larger cohorts and multi-target stimulation protocols are warranted to refine the diagnostic and prognostic value of TMS–EEG and to explore its potential role in guiding individualized neuromodulation strategies for PPA.

## Data Availability

All data produced in the present study are available upon reasonable request to the authors

## Acknowledgements

Data used in preparation of voxel-based morphometry (VBM) analysis for this article were obtained from the Alzheimer’s Disease Neuroimaging Initiative (ADNI) database (adni.loni.usc.edu). As such, the investigators within the ADNI contributed to the design and implementation of ADNI and/or provided data but did not participate in analysis or writing of this report. A complete listing of ADNI investigators can be found at: http://adni.loni.usc.edu/wp-content/uploads/how_to_apply/ADNI_Acknowledgement_List.pdf

Data collection and sharing for this project was funded by the Alzheimer’s Disease Neuroimaging Initiative (ADNI) (National Institutes of Health Grant U01 AG024904) and DOD ADNI (Department of Defense award number W81XWH-12-2-0012). ADNI is funded by the National Institute on Aging, the National Institute of Biomedical Imaging and Bioengineering, and through generous contributions from the following: AbbVie, Alzheimer’s Association; Alzheimer’s Drug Discovery Foundation; Araclon Biotech; BioClinica, Inc.; Biogen; Bristol-Myers Squibb Company; CereSpir, Inc.; Cogstate; Eisai Inc.; Elan Pharmaceuticals, Inc.; Eli Lilly and Company; EuroImmun; F. Hoffmann-La Roche Ltd and its affiliated company Genentech, Inc.; Fujirebio; GE Healthcare; IXICO Ltd.; Janssen Alzheimer Immunotherapy Research & Development, LLC.; Johnson & Johnson Pharmaceutical Research & Development LLC.; Lumosity; Lundbeck; Merck & Co., Inc.; Meso Scale Diagnostics, LLC.; NeuroRx Research; Neurotrack Technologies; Novartis Pharmaceuticals Corporation; Pfizer Inc.; Piramal Imaging; Servier; Takeda Pharmaceutical Company; and Transition Therapeutics. The Canadian Institutes of Health Research is providing funds to support ADNI clinical sites in Canada. Private sector contributions are facilitated by the Foundation for the National Institutes of Health (www.fnih.org). The grantee organization is the Northern California Institute for Research and Education, and the study is coordinated by the Alzheimer’s Therapeutic Research Institute at the University of Southern California. ADNI data are disseminated by the Laboratory for Neuro Imaging at the University of Southern California.

This work was supported by the Italian Ministry of University and Research (MUR) under the “Tuscany- Health Ecosystem” project (code ECS00000017; CUP B63C2200068007), within Italy’s National Recovery and Resilience Plan (PNRR), Mission 4 – Component 2 – Investment 1.5, funded by the European Union – NextGenerationEU.

---

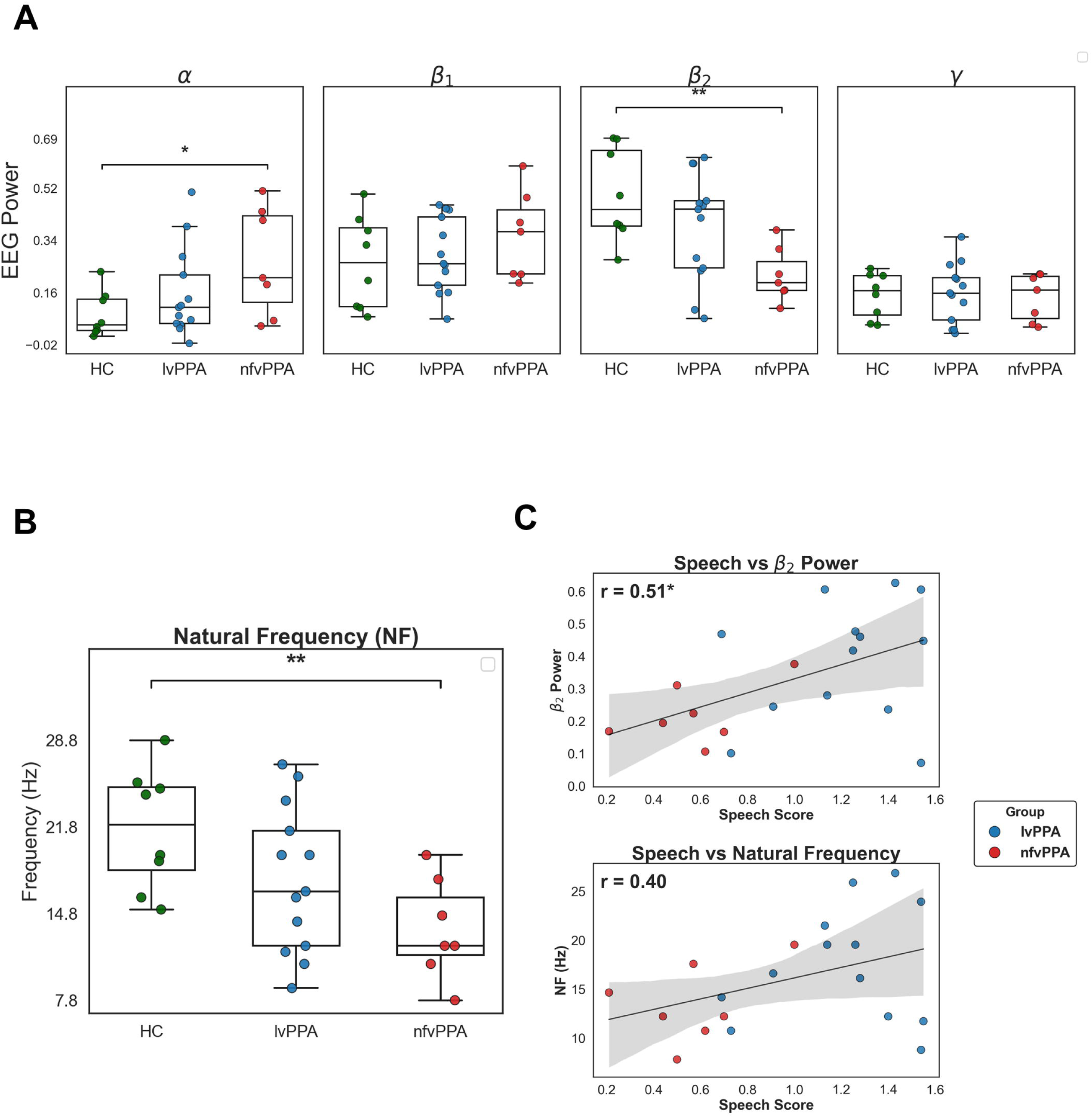

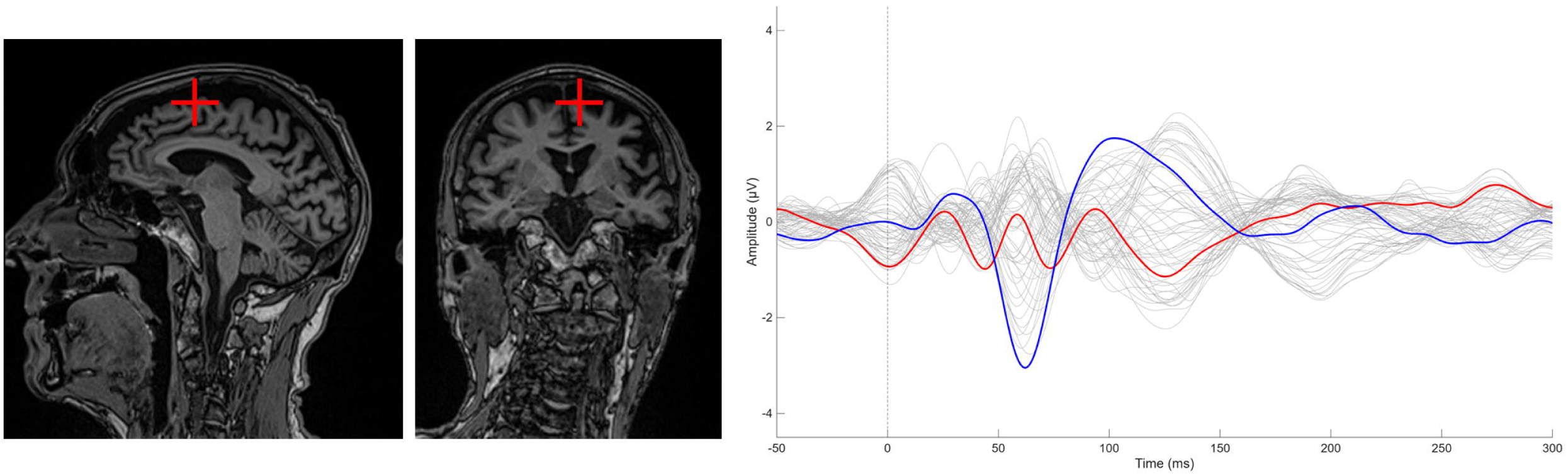

