## Supplementary materials for "TMS–EEG Reveals Distinct Cortical Signatures in Non-Fluent PPA"

**Title:**

**Running title:** TMS–EEG Biomarkers in nfvPPA

**Supplementary Table 1.** Comparison of stimulation parameters, induced E-field, target coordinates, and TMS-evoked potentials across participant groups: nfvPPA (n = 7), lvPPA (n = 13), and healthy controls (HC, n = 8). Values are mean (SD). No significant differences were observed between groups. Electrode of interest (EOI) indicates the electrodes used for main analyses.

|  | **nfvPPA (*n* = 7)** | **lvPPA (*n* = 13)** | **HC (*n* = 8)** |  |
| --- | --- | --- | --- | --- |
| Stimulation intensity, % stimulator output, mean (SD) | 62.1 (7.88) | 60.4 (11.8) | 55.9 (5.89) | *P* = 0.435 |
| Maximum induced E-field, V/m, mean (SD)* | 113.1 (22.8) | 100.2 (21.2) | 84.1 (24.8) | *P* = 0.062 |
| MNI target coordinates (x), mean (SD) | -15.7 (7.99) | -15.7 (5.64) | -15.8 (5.43) | *P* = 0.999 |
| MNI target coordinates (y), mean (SD) | 10.5 (5.22) | 15.2 (10.02) | 12.2 (16.8) | *P* = 0.342 |
| MNI target coordinates (z), mean (SD) | 63.1 (8.13) | 63.3 (7.97) | 68.5 (6.23) | *P* = 0.267 |
| Electrode of interest (EOI) | 3 FC1, 1 FCZ, 1 C1, 2 C3 | 5 FC1, 1 FCZ, 1 FC2, 1 FZ, 1 CZ, 2 C1, 2 C3 | 5 FC1, 2 FCZ, 1 F2 | *P* = 0.686 |
| Peak-to-peak over the EOI, μV, mean (SD) | 7.86 (2.25) | 8.95 (2.60) | 9.88 (4.03) | *P* = 0.440 |
| Peak-to-peak over FC1, μV, mean (SD) | 5.33 (3.62) | 7.79 (3.15) | 9.43 (4.39) | *P* = 0.145 |

nfvPPA = non-fluent variant PPA, lvPPA = logopenic variant PPA, HC = healthy controls, MMSE = Mini Mental State Examination, SD = standard deviation.

*The maximum induced E-field values represent the 95^th^ percentile over a ROI centered over -24, 4, 67 MNI coordinates, corresponding to upper dorsal border of left PMd^73^.

**Supplementary Table 2.** Left vs. right hemisphere comparison of PMd target coordinates, induced E-field, and peak-to-peak values in a subsample of participants (n = 9) for whom right hemisphere data were available. No significant differences were observed between hemispheres.

|  | **Left** | **Right** |  |
| --- | --- | --- | --- |
| Maximum induced E-field*, V/m, mean (SD) | 94.21 (20.2) | 90.26 (25.2) | *P* = 0.437 |
| MNI target coordinates (x), mean (SD) | -12.6** (6.87) | 16.4 (5.09) | *P* = 0.055 |
| MNI target coordinates (y), mean (SD) | 15.7 (9.41) | 17.9 (10.3) | *P* = 0.309 |
| MNI target coordinates (z), mean (SD) | 64.7 (4.00) | 61.6 (5.57) | *P* = 0.301 |
| Peak-to-peak over the EOI, μV, mean (SD) | 9.36 (2.57) | 7.94 (3.70) | *P* = 0.174 |

* The maximum induced E-field values represent the 95th percentile over a ROI centered over -24, 4, 67 MNI coordinates, corresponding to upper dorsal border of left PMd^73^. We used homologous coordinates (24, 4, 67) to define the right ROI.

**Given that MNI x-coordinates are negative in the left hemisphere and positive in the right, we compared the absolute x-values to assess lateral distance from the midline.

**TMS–EEG results on FC1 electrode**

The TMS–EEG results over the FC1 channel are reported in Supplementary Fig. 1. When considering the same electrode (i.e., FC1) for all the subjects, we observed a significant effect of group for the high-beta range (*F*(2,25) = 5.50, *P* = 0.008); non-fluent patients were characterized by lower power levels in that specific frequency band compared with healthy controls (*Tukey-corrected* *P* = 0.016). No group differences were present in the alpha (*F*(2,25) = 2.94, *P* = 0.072), low-beta (*F*(2,25) = 0.84, *P* = 0.443) and gamma band (*F*(2,25) = 0.06, *P* = 0.940).

We also observed lower natural frequency in nfvPPA compared to controls (*F*(2,25) = 4.52, *P* = 0.021; *Tukey-corrected* *P* = 0.016). Speech rate significantly correlated high-beta (*r* = 0.51, *P* = 0.022) but not with natural frequency (*r* = 0.40, *P* = 0.081).

**Figure legends**

**Supplementary Figure 1 TMS–EEG spectral measures at a fixed electrode (FC1).** (**A**) Box-plots of evoked spectral power in each frequency band for each group. (**B**) Box-plots of natural frequency values for each group. (**C**) Linear relations between the speech rate at clinical evaluation and each TMS–EEG outcome: evoked high-beta power (above) and natural frequency (below). Asterisks indicate significant group differences or correlations (*P* < 0.05; *P* < 0.01; *P* < 0.001).

**Supplementary Figure 2** **Anatomical location of stimulation and grand-average EEG in a severely atrophic nfvPPA subject.** **(A)** Sagittal and coronal T1-weighted MRI slices showing the site of stimulation. **(B)** Butterfly plot of the grand-average EEG across all channels. The channel with the maximal peak-to-peak amplitude (CP3) is highlighted in blue, and FC1 is highlighted in red. Early peak-to-peak amplitudes remain relatively small (< 4 µV), illustrating why this subject was excluded from the main analysis.
